# Humoral immune response and prolonged PCR positivity in a cohort of 1343 SARS-CoV 2 patients in the New York City region

**DOI:** 10.1101/2020.04.30.20085613

**Authors:** Ania Wajnberg, Mayce Mansour, Emily Leven, Nicole M. Bouvier, Gopi Patel, Adolfo Firpo, Rao Mendu, Jeffrey Jhang, Suzanne Arinsburg, Melissa Gitman, Jane Houldsworth, Ian Baine, Viviana Simon, Judith Aberg, Florian Krammer, David Reich, Carlos Cordon-Cardo

## Abstract

**Background:** Severe acute respiratory syndrome coronavirus 2 (SARS-CoV-2) has caused a global pandemic. The percentage of infected individuals who seroconvert is still an open question. In addition, it has been shown in some individuals that viral genome can still be detected at considerable time post symptom resolution. Here we investigated both seroconversion and PCR-positivity in a large cohort of convalescent serum donors in New York City.

**Methods:** Individuals with confirmed or suspected SARS-CoV-2 infection were screened via PCR for presence of viral genome and via enzyme-linked immunosorbent assay for presence of anti SARS-CoV-2 spike antibodies.

**Results:** All but three confirmed SARS-CoV-2 patients seroconverted to the SARS-CoV-2 spike while only 37.4% of suspected SARS-CoV-2 patients seroconverted. PCR-positivity was detected up to 28 days from symptom resolution.

**Conclusions:** Here we show that the vast majority of confirmed COVID19 patients seroconvert, potentially providing immunity to reinfection. We also report that in a large proportion of individuals, viral genome can be detected via PCR in the upper respiratory tract for weeks post symptom resolution, but it is unclear if this signal represents infectious virus.

## Introduction

Severe acute respiratory syndrome coronavirus 2 (SARS-CoV-2) has rapidly spread around the world, leading to unprecedented strain on health care systems and economies, causing over 290,000 infections and 17,000 deaths in New York State (source, New York State, Department of Health as of April 28^th^ 2020) at the time of writing this report. Significant disruptions to daily life have been enacted in order to flatten the epidemic curve. In order to avoid spread of SARS-CoV-2 and to help standardize definitions of clearance, it is important to understand the duration of SARS-CoV-2 nucleic acids within the nasopharynx and the time course to the mounting of an antibody response to this new viral pathogen.

Current US Centers for Disease Control and Prevention (CDC) guidelines suggest that people with confirmed or suspected SARS-CoV-2 infection should remain in isolation for at least seven days from symptom onset and return to work if they have been asymptomatic for 72 hours. However, to date, there are limited data that help to define the time to viral clearance from illness onset and cessation of symptoms. A prior case study suggested that SARS-CoV-2 can exhibit ongoing viral shedding for a median of 2.5 days (range 1–8 days) after complete symptom resolution, but it remains unclear whether this viral shedding poses a risk for forward transmission.^1^ A small case sample of four recently hospitalized patients found that they were SARS-CoV-2 positive on repeat polymerase chain reaction (PCR) testing five to 13 days after discharge.^2^ Other studies have found viral shedding for up to six weeks after symptoms.^3^ From work with the 2003 SARS-CoV-1it is not clear if detection of viral genome of this duration indicates prolonged infectivity or presence of nonviable virus.^4^ A clearer understanding of duration of viral shedding is critical to preventing transmission by infected individuals, particularly as they begin to feel well enough to resume normal activities. Understanding time to PCR clearance may also help guide isolation durations and return to work clearance, as well as clarify the utility of negative PCR testing as part of defining disease clearance.

There is limited data worldwide on the development of antibodies to SARS-CoV-2, particularly the formation of IgG. While there is concern regarding efficacy of antibody testing for diagnosis of SARS-CoV-2, little is known about long lasting immunity. One study measured neutralizing antibodies in 175 hospitalized patients and found that 64% had high antibody titers, 30% had weak antibody response, and 6% had undetectable titers.^5^ Studying the plasma from previously infected individuals may further our understanding of the timing and strength of different populations’ antibody response to this novel illness, delineate duration of antibody presence, and identify cases of possible reinfection. Additionally, individuals with high antibody titers may become donors for convalescent plasma treatment for critically ill patients as part of ongoing studies of this therapeutic option.^6–8^

Here we present a large data set of serum antibody testing in persons who have fully recovered after mild illness from SARS-CoV-2 at Mount Sinai Hospital. Our aim is to describe the time to SARS-CoV-2 PCR clearance from the nasopharynx, the rates of IgG development, and time to serum IgG development from onset and resolution of symptoms in 1,343 participants with prior confirmed or suspected SARS-CoV-2 infection.

## Methods

We conducted an outreach program in the New York City area, including parts of New York State, Connecticut, and New Jersey, to identify people recovered from SARS-CoV-2 for nasopharyngeal PCR (cobas® SARS-CoV-2, Roche Diagnostics, Indiana) and serum IgG titer measurement (ELISA Mount Sinai).^9,10^ Participants were tested between March 26 and April 10, 2020. We recruited participants via REDCap^®^ (Vanderbilt University, Tennessee) online survey response which was advertised on our hospital website, and subsequently shared by multiple news organizations and public officials in New York. REDCap respondents were deemed eligible if they had previously tested positive for SARS-CoV-2, or if they were symptomatic with suspected SARS-CoV-2 and lived with someone with a positive SARS-CoV-2 PCR test, had been told by a physician that they had symptoms consistent with SARS-CoV-2, or were healthcare workers. We only included participants who self-reported suspicious symptoms after February 1^st^ 2020, as this is when it is believed SARS-CoV-2 began to spread in New York City. Additionally, only participants who were asymptomatic at time of survey were outreached. Respondents self-reported date of symptom onset, date of positive SARS-CoV-2 test (if applicable), and last date of symptoms. Duration of symptoms was calculated from these self-reported dates.

During the first two weeks of the survey, we tested for SARS-CoV-2 in the nasopharynx by PCR as well as IgG antibody (Ab) in the serum of every individual while in the third week the testing was limited to SARS-CoV-2 antibodies only. The rationale for this change was that we had a far larger testing volume combined with the fact that many of the participants reported symptom resolution >14 days prior to testing. During week one, participants were brought in ten days after they had a confirmed/suspected diagnosis and had been asymptomatic for at least three days. In week 2, as we identified more potential donors and learned more about our antibody assay, we extended our timeline to 14 days after symptoms onset, with at least three days asymptomatic. In week 3, we included participants 21 days or more after symptom onset, who had been completely asymptomatic for at least 14 days.

SARS-CoV-2 PCR was considered positive if detected on nasopharyngeal swab. We measured serum IgG antibody titers using a serologic enzyme-linked immunosorbent assay (ELISA) developed at Icahn School of Medicine at Mount Sinai and described on March 18, 2020; this serum test has a sensitivity of 92% and a specificity >99%. Serum IgG titers were considered “strongly positive” if they were detected at titers of 1:320 or higher (highest dilutions were 1:320, 1:960, 1:2880), and considered “weakly positive” if detected at titers of 1:80 and 1:160. Negative was defined as titers below 1:80 (and is shown in figures as 1:40). All interested participants with antibody titers >1:320 and negative SARS-CoV-2 PCR swabs were screened by the New York Blood Center using standard criteria for plasma donation and included as donors in our convalescent plasma study if eligible as per CFR Title 21. Participants with weakly positive antibody titers were invited to return for repeat serum titer testing at least seven days after their initial antibody test. Participants with positive PCR swabs and antibodies were asked to return for PCR testing at least three days after initial PCR test so they could be referred for plasma donation once the virus had fully cleared. Given the frequent serial detection of SARS-CoV-2 by nasopharyngeal PCR using three-day increments, on April 3^rd^, 2020, we began to ask participants to return at least ten days after the last positive PCR for re-testing.

One-way ANOVA and Fisher’s exact test were used to measure the association of age, gender, symptom duration, and days from symptom onset and resolution with positive antibody results. This study was reviewed and approved by our institutional review board.

## Results

We measured SARS-CoV-2 antibody titers in 1,343 people over the first three weeks of the survey (March 26, 2020 to April 10, 2020) using a now FDA approved two step ELISA.^9,10^ The average age of the participants was 40 years (range 17 to 76 years) with 256 (19%) between the ages of 17 and 29, 968 (72%) age 30–59, and 119 (9%) 60 or older. Fifty three percent were male and 47% had confirmed SARS-CoV-2 diagnosis by prior PCR testing. Median days between symptom onset to serum antibody test was 24 days (range 3-70), median days between symptom resolution to antibody test was 15 (4–77) and median duration of symptoms was 9 days (1–67). Of the 1,343 total participants, almost all were outpatients who had experienced mild to moderate symptoms, and only 3% were seen in our emergency department or hospital.

Seven hundred and sixty-one (57%) participants of the total sample were antibody positive, 61 (5%) were weakly positive, and 521 (39%) were negative. Neither age, gender, nor symptom duration was associated with antibody response. In the 584 participants for whom both nasopharyngeal PCR testing and serum antibody testing was available, SARS-CoV-2 RNA was detected in 249 (42%) at a median of 20 days from symptom onset (range 11–43) and 12 days from symptom resolution (range 5–28).

Six hundred and twenty-four participants had confirmed SARS-CoV-2 disease by PCR prior to coming for testing, either by self-report or documented in our electronic medical record; if self-reported, participants provided the date of testing. In this subgroup the average age was 39 (range 17–76) and 59% were male. At first test, five hundred and eleven (82%) were strongly antibody positive at titer >= 1:320, 42 (7%) were weakly positive, and 71 (11%) were negative (**Figure 1A**). We asked the 18% with initial negative or weakly positive antibody response to return for a second test ten or more days later. At the time of the first test, 217 (35%) were still PCR positive (range 5–22 days from symptom resolution) (**Figure 2**). Median duration of symptoms in this group was nine days (range 1–31). Neither age nor sex was associated with a strong antibody response. Greater number of days between symptom onset and antibody test was associated with a higher titer antibody test (23 days versus 20 days). Symptom duration was also associated with higher antibody titers (9 days versus 7 days). (**Table 2**)

**Figure 1:**
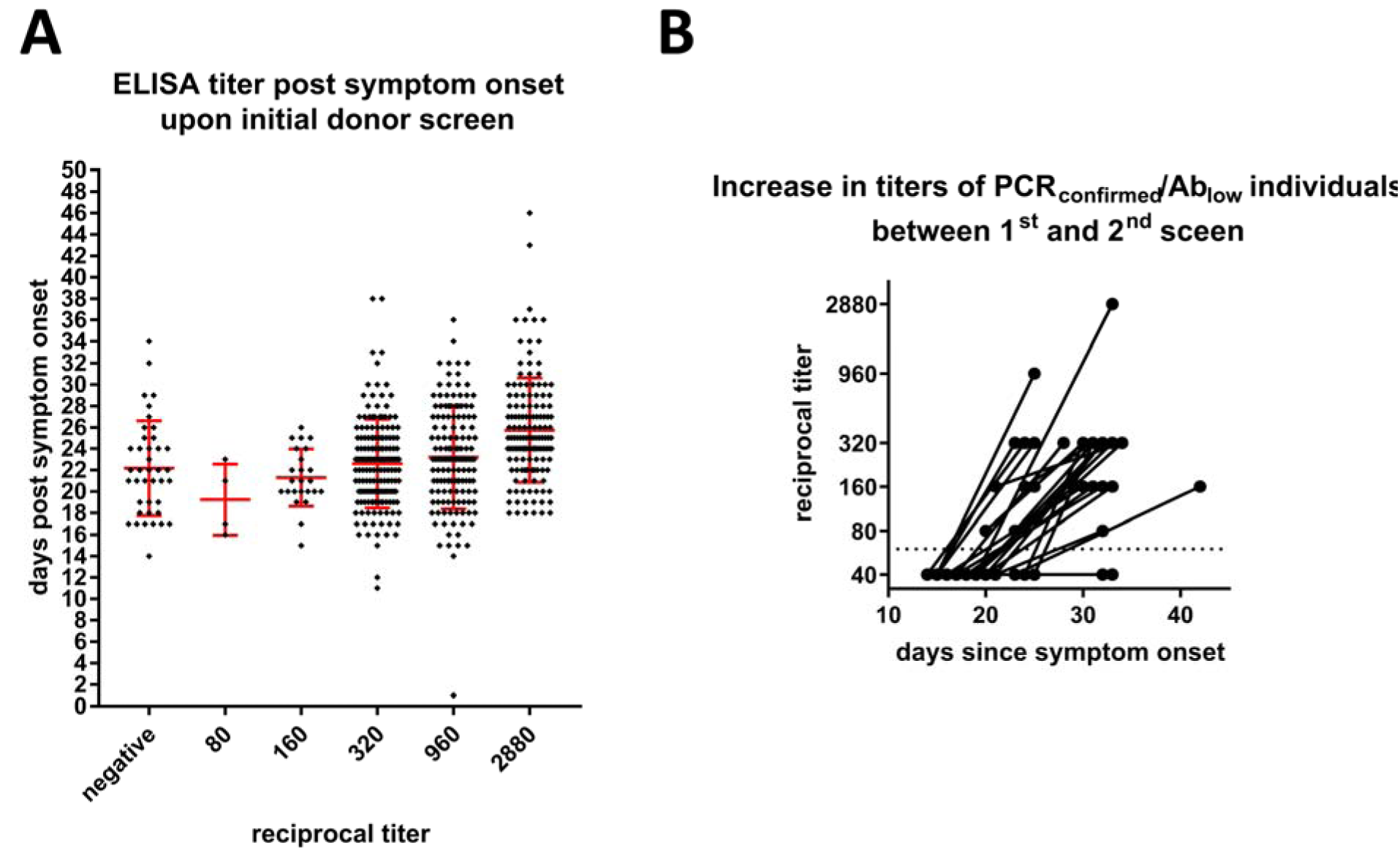
Antibody responses in PCR-confirmed COVID19 individuals. **A** shows antibody testing results days post symptom onset stratified into titer categories. The highest titer category contains both 1:2880 and >1:2880 titers. Bars represent the mean; error bars the standard deviation. **B** Individuals with negative titers were recalled for retesting. Both the original test result and the second test result post day onset are shown. Negative titers were assigned a value of 1:40 for representation purpose, the line represents the cut-off between positive and negative titers. Only results for individuals for which date of symptom onset is known are shown.

**Figure 2:**
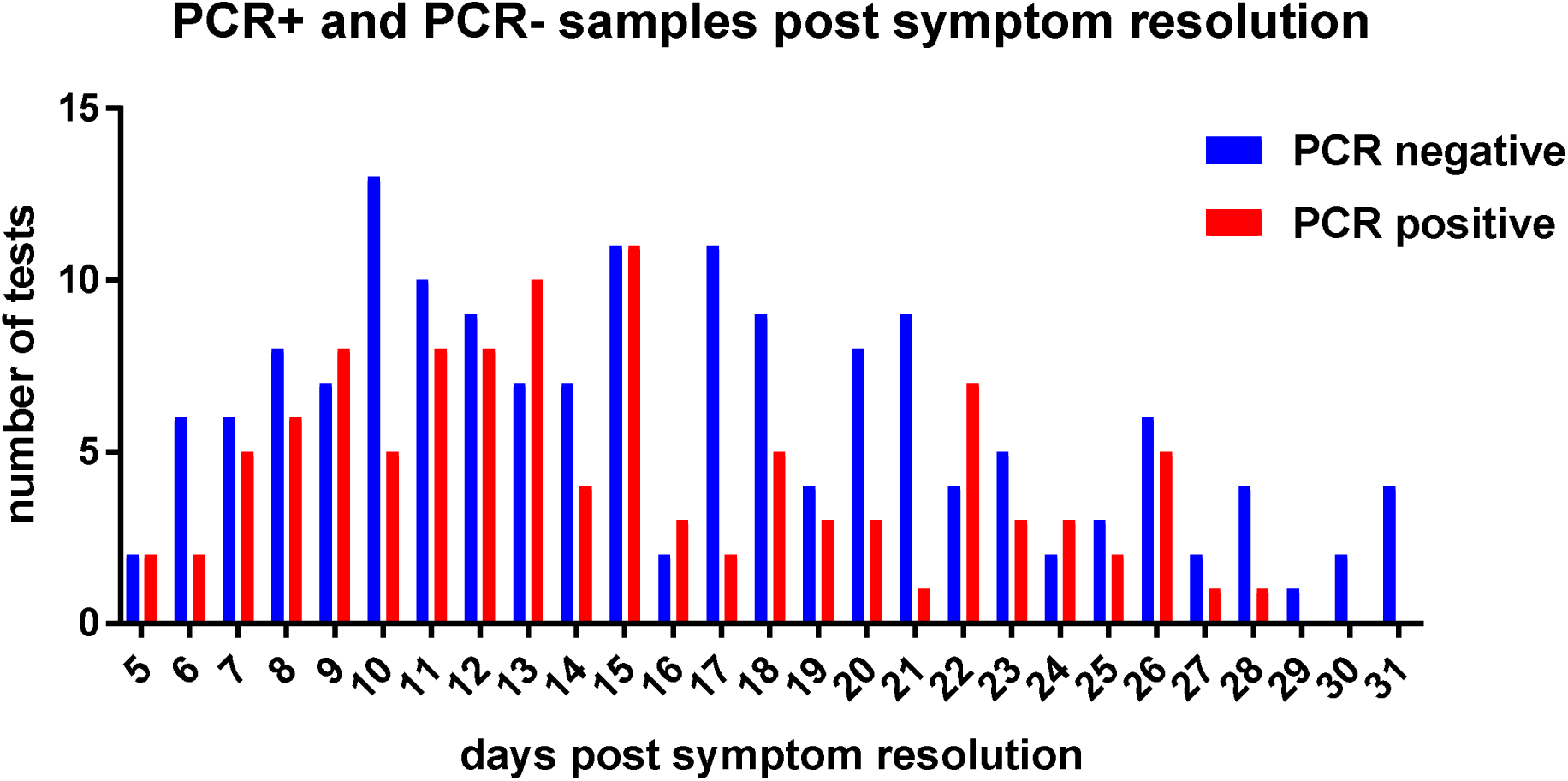
PCR results of initially PCR-confirmed COVID19 individuals. Viral genome was detected in nasopharyngeal swaps of individuals screened as plasma donors. Number of subjects positive or negative days post symptom resolution are shown. More than one result from an individual might be shown if tested more than once on different days. Only results for subject for which a date of symptom resolution was available are shown.

In the subgroup of 719 participants with suspected disease that did not have confirmed SARS-CoV-2 infection, average age was 41 (17–76) and 47% were male. 250 (35%) were strongly antibody positive, 19 (3%) were weakly positive, and 436 (62%) were negative. Antibodies were measured a median of 28 days from symptom-onset and 19 days from date of symptom resolution. In this group, neither age, gender, nor symptom duration was associated with stronger antibody response. (**Table 1)**

**Table 1.**
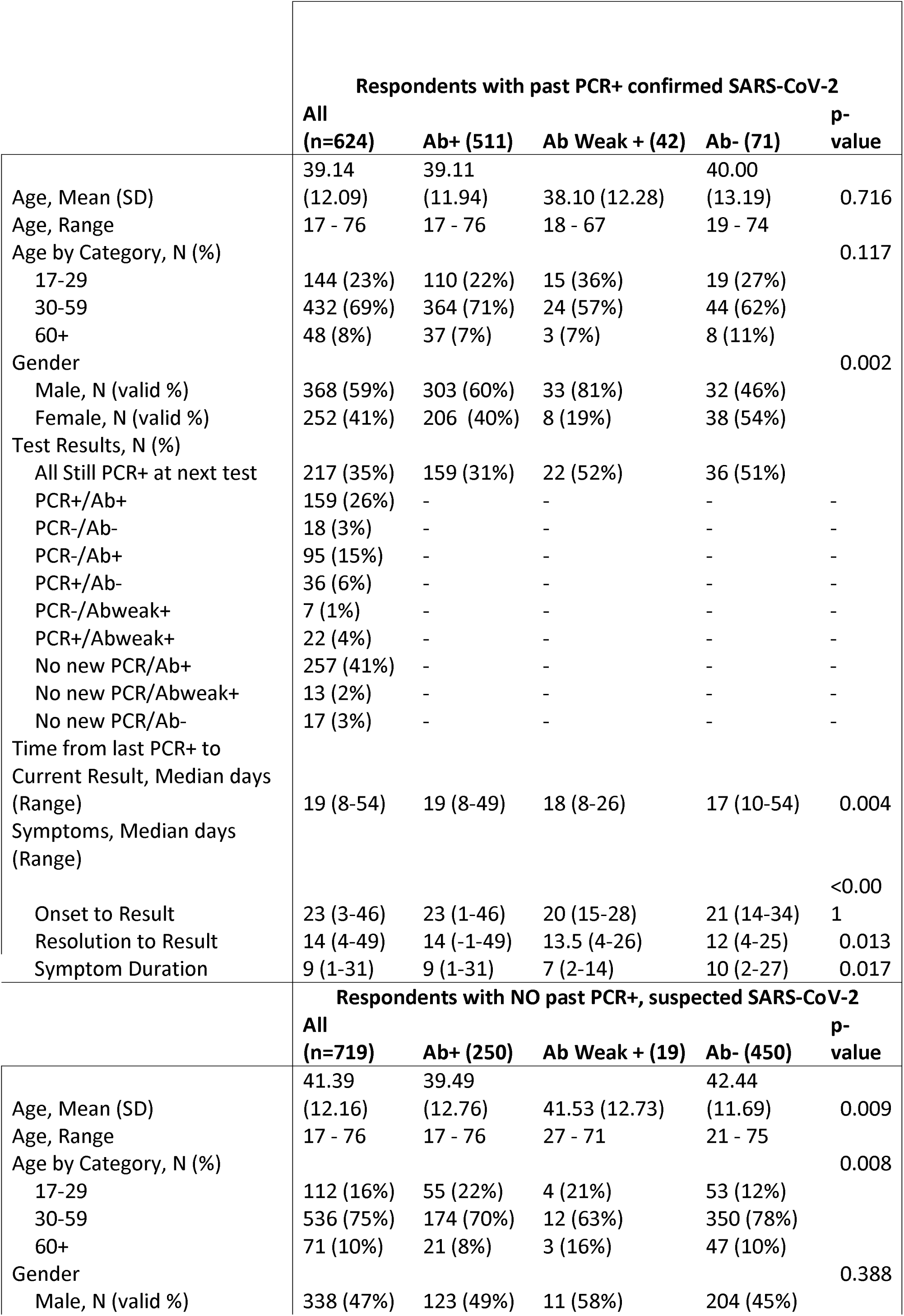
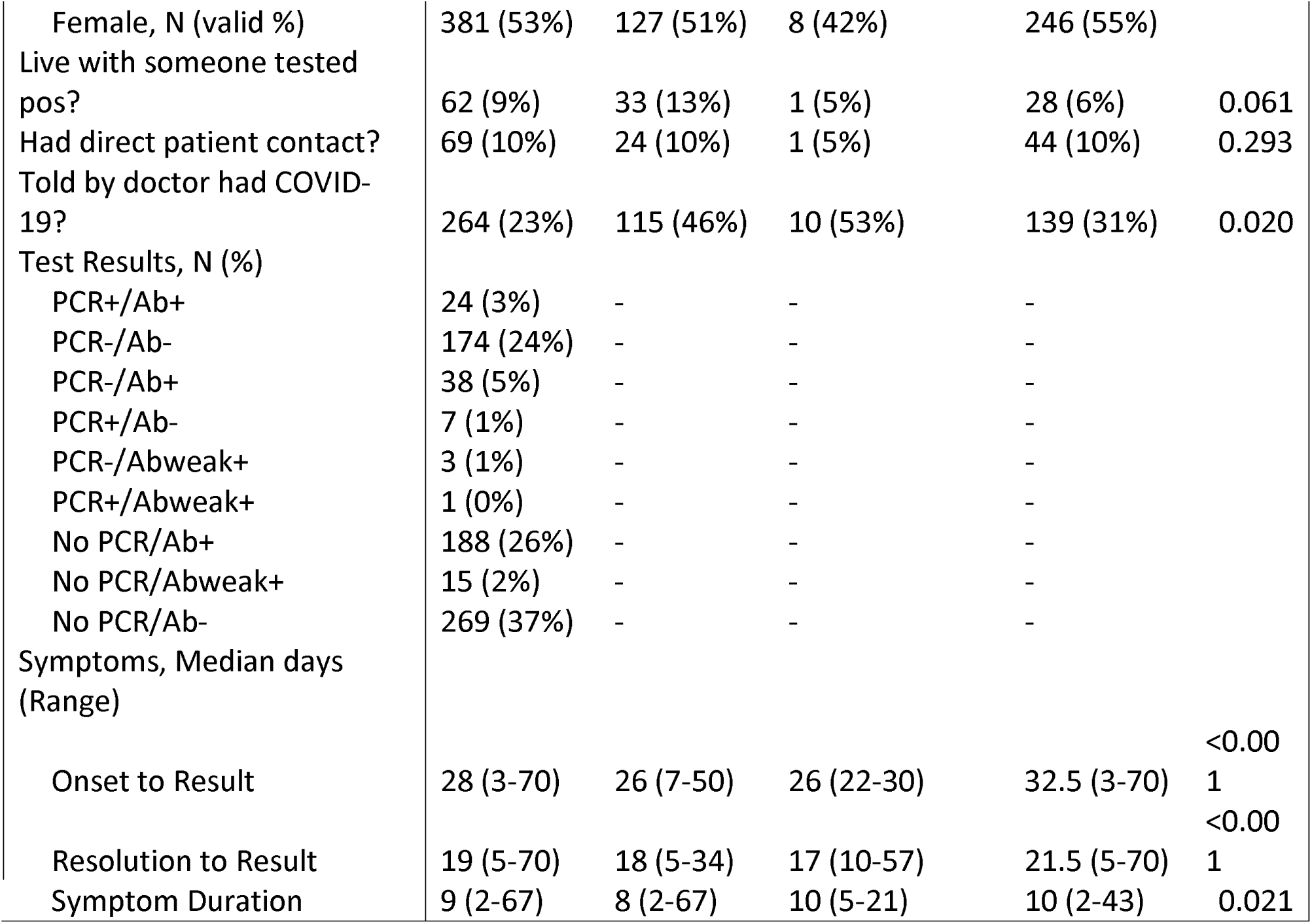

Of the 113 participants with PCR confirmed SARS-CoV-2 and weakly positive or negative titers on their first serum antibody test, 64 have returned for follow up antibody titers at the time of submission. Of these, 57 (89%) displayed increased titers between the two tests, a median of 13 days (5–25) later (**Figure 1B**). Four remained weakly positive, and three remained negative. The three that remained negative all self-reported positive PCR testing (none were documented in our EMR). (**Table 2)**

**Table 2.**
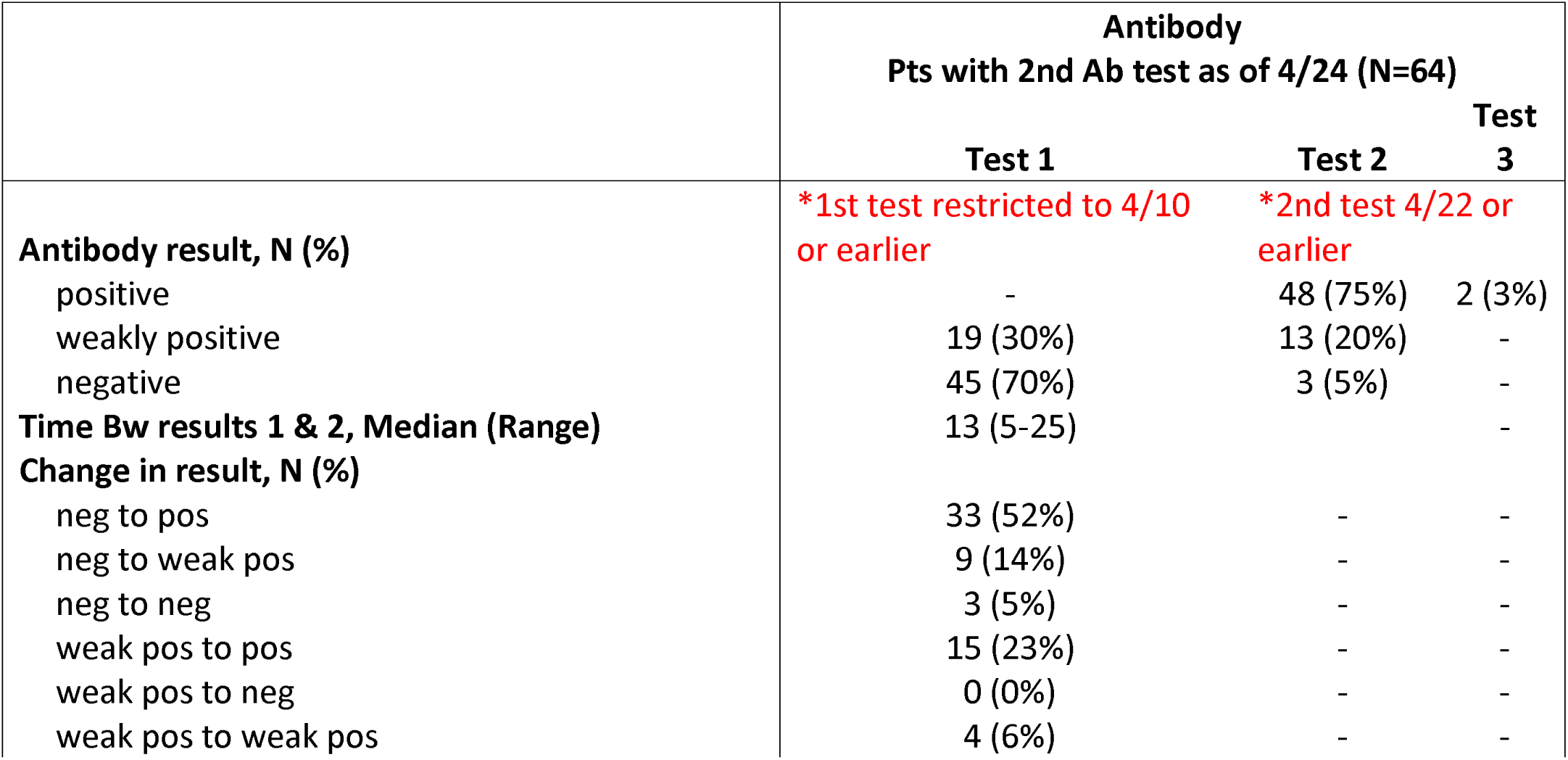
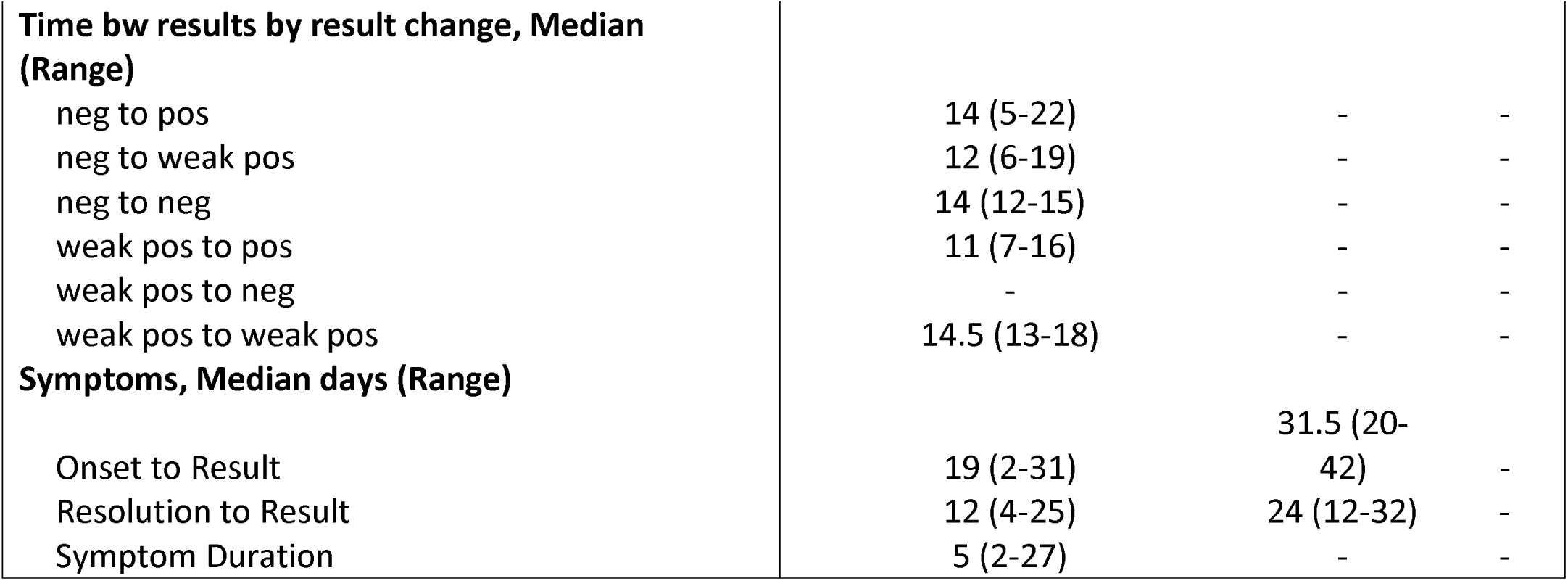

While all survey participants self-reported complete resolution of symptoms three to 14 or more days prior to testing, 249 (19%) tested positive for nasopharyngeal SARS-CoV-2 RNA. The maximum time of positive nasopharyngeal PCR testing was 43 days from symptom onset and 28 days from symptom resolution. For the 182 individuals who returned for repeat nasopharyngeal swabbing at least three days after prior positive test, 61% were negative on the repeat test, a median of 10 days (range 3–21) after the first test. Seventy (39%) remained positive and were rescheduled for another nasopharyngeal PCR 7–10 days later. (**Table 3**).

**Table 3.**

|  | **PCR** |  |  |
| --- | --- | --- | --- |
|  | **Pts with 2nd PCR test as of 4/24 (N=182)** |  |  |
|  | **Test 1** | **Test 2** | **Test 3** |
| **PCR result, N (%)** | **\*1st test restricted to 4/10 or earlier** | **\*2nd test 4/22 or earlier** |  |
| Detected | 182 (100%) | 70 (39%) | 8 (57%) |
| Not Detected | - | 112 (61%) | 6 (43%) |
| **Time Bw results 1 and 2, Median (Range)** | 10 (3-21) | - | - |
| **Change in result, N (%)** |  |  |  |
| Detected to Detected | 70 (39%) | - | - |
| Detected to Not Detected | 12 (61%) | - | - |
| **Time bw results by result change, Median (Range)** |  |  |  |
| Detected to Detected | 10 (3-21) | - | - |
| Detected to Not Detected | 10 (4-21) | - | - |
| **Symptoms, Median days (Range)** |  |  |  |
| Onset to Result | 20 (1-43) | 30 (6-50) | - |
| Resolution to Result | 12 (-1-20) | 22 (4-34) | - |
| Symptom Duration | 8 (1-27) | - | - |

## Discussion

Our survey provides a large cross-sectional representation of SARS-CoV2 RNA and antibodies found in participants recruited after recovery from SARS-CoV-2 during the early weeks of the outbreak in New York City. Understanding the duration of potential infectiousness and the time to IgG antibody response are critical to the containment of SARS-CoV-2 spread, and the plans for widespread antibody testing over the coming months. Some countries, states and organizations may even be considering antibody testing prior to letting individuals return to work.

In contrast to some of the prior literature on formation of antibodies, over 99% of the patients who self-reported or had laboratory documented SARS-CoV-2 infection developed IgG antibodies using our assay. Additionally, our findings suggest that IgG antibodies develop over a period of 7 to 50 days from symptom onset and 5 to 49 from symptom resolution, with a median of 24 days from symptom onset to higher antibody titers, and a median of 15 days from symptom resolution to higher antibody titers. This suggests that the optimal time frame for widespread antibody testing is at least three to four weeks after symptom onset and at least two weeks after symptom resolution. In our survey, we did not find evidence for a decrease in IgG antibody titer levels on repeat sampling.

Although we do not yet know what, if any, immunity is conferred by IgG or the duration of the IgG response, at this time it seems likely that IgG to SARS-CoV-2 may confer some level of immunity based on what is known about viral immunity to other pathogens. In prior studies of SARS-CoV-1 and Middle East Respiratory Syndrome patients, IgG peaked within months of primary infection and waned over time.^11–13^ Similar observations have been made with human coronaviruses were immunity can confer at least limited protection.^13^ In order to study the duration of IgG antibody response to SARS-CoV-2, we plan to follow our cohort for the next six months in order to track titer levels.

Among participants who did not have prior PCR but who were deemed high risk, i.e., people with symptoms consistent with SARS-CoV-2 who were told by a healthcare provider they had presumed infection, lived with someone with confirmed infection, or were healthcare workers themselves, we found 36% of this population had IgG antibodies to SARS-CoV-2. This finding suggests that a majority of participants suspected of having Covid-19 actually were not infected with SARS-CoV-2; however, it may also include a false negative rate of our assay (which has a 92% sensitivity) or insufficient time for participants to mount an IgG antibody response. This underscores the importance of expanded PCR testing in order to improve diagnosis of this disease even in minimally symptomatic individuals.

The 19% of participants who remained PCR positive despite self-reporting full resolution of symptoms bring to light important considerations regarding the possible duration of viral transmission, and the limited utility of PCR testing to ensure clearance. This positive PCR finding could represent shedding of nonviable virus, non-infectious genome fragments or viruses engulfed by immune cells, asymptomatic carriers of SARS-CoV-2, or ongoing disease despite full resolution of symptoms. Detection of viral genome even months after resolution of infection has been shown for viruses like measles virus.^14^ Further studies are warranted to determine if nasopharyngeal PCR positivity is related to transmission, and if so, for how long. This will have significant implications in terms of guidance of when recovered SARS-CoV-2 individuals should end self-isolation, currently recommended at least seven days after symptom onset with at least 72 hours without fever off of antipyretics. If PCR positivity is a result of identifying noninfectious genome or nonviable virus, it may be necessary to avoid use of PCR as a definition of clearance in SARS-CoV-2.

There are limitations to our evaluation. All participants had mild disease, and thus these data may not reflect PCR or Ab findings in a moderately or severely ill population. Participants were recruited based on self-referral and self-reporting, which may have led to recall bias in terms of dates of symptom onset, resolution and duration, and may have led us to miss asymptomatic carriers who did not inquire about testing. Additionally, given recruitment via an English-language online survey and our use of a single collection site, our sample likely included more recovered participants of younger ages with internet access and the ability to travel to our site for testing. Furthermore, we did not collect rigorous data regarding symptom severity which could potentially be related to the timeline and strength of IgG antibody response to SARS-CoV-2.

Future studies are planned to help us understand the magnitude and duration of the IgG response in patients recovered from SARS-CoV-2, and what antibody titer may be necessary to protected individuals from reinfection. We also hope to better understand which, if any, patients do not mount an IgG immune response. Finally, the clinical significance of prolonged positive SARS-CoV-2 nasopharyngeal PCR in the absence of symptoms requires further clarification.

## Conclusion

Duration of nasopharyngeal PCR SARS-CoV-2 PCR detection and time to mount IgG antibody response have important implications for the spread of this virus and risk for re-infection among individuals. In our sample, we found that 19% of people continue to have nasopharyngeal PCR positivity two or more weeks after symptom resolution, and that it takes three or more weeks to mount an IgG antibody response believed to be potentially protective against future infection. Reassuringly, we found that almost all participants with confirmed SARS-CoV-2 infection in our study mounted an IgG immune response to this disease. Taken together, these findings will be pivotal in understanding disease activity of SARS-CoV-2 moving forward.

## Data Availability

Authors are responsible for all data presented in the manuscript.

## Acknowledgements

Thank you to all the patients who generously came in for testing and plasma donation. Thanks to Kim Stone and Kim Muellers for their work on data analysis.

Work on SARS-CoV-2 immunity in the Krammer is supported by the National Institute of Allergy and Infectious Disease (NIAID) Collaborative Influenza Vaccine Innovation Centers (CIVIC) contract 75N93019C00051 and the NIAID Centers of Excellence for Influenza Research and Surveillance (CEIRS) contract HHSN272201400008C as well as institutional and philanthropic funding.

